# Stool Antigen Test is Effective and Sensitive for Detecting *Helicobacter Pylori* Infection in Bangladeshi Peptic Ulcer Patients

**DOI:** 10.1101/2021.10.22.21265367

**Authors:** Afreen Sultana, Shakeel Ahmed, Ershad Uddin Ahmed, Abul Faisal MD. Nuruddin Chowdhury, Abul Kalam, Arifur Rahman, Farhana Akter, A. H. M. Saiful Karim Chowdhury, Sabrina Sharmin, Jannatul Ferdous Mustry, Ayan Saha

## Abstract

**Aims:** *Helicobacter pylori* (*H. pylori*) is the primary causative agent of peptic ulcer in multiple developing countries, including Bangladesh. This study was designed to investigate the diagnostic value of a rapid immunochromatography-based *H. Pylori* stool antigen (HpSAg) test to screen *H. pylori* infection in Bangladeshi population.

**Methodology and results:** A total of 140 suspected peptic ulcer patients who underwent upper gastrointestinal endoscopy at Chittagong Medical College and Hospital, Chattogram, Bangladesh, were included in the study. Histopathology, Rapid Urease Test (RUT), and Microscopic examination of the stained smears were conducted to define *H. pylori* positive cases. Later, stool antigen detection test was done in H. *pylori* positive status group, *H. pylori* negative status group, indeterminate status group, and healthy controls. Out of 140 peptic ulcer suspected patients, 75 (53.6%) patients were confirmed to have peptic ulcer or erosions. Although the proportion of antral erosion was 57.4% in patients who were below 40 years, the proportion decreased to 23.4% in patients over 40 years. Patients over 40 years were mostly suffering from Prepyloric erosion (42.9%). All peptic ulcer patients were also positive on histopathological analysis. However, micorscopic grading of curved bacilli and RUT found 93.3% (70/75) and 89.3% (67/75) patients positive, respectively. High sensitivity (95%), specificity (80%), and diagnostic accuracy (91%) scores for HpSAg assay was obtained in our study.

**Conclusions, significance and impact of studies:** The HpSAg test, for a comparatively less sophisticated assay, can be efficient in detecting the presence of *H. pylori* pre-and post-therapy and provide more valid test results than other invasive test methods.

## 1. INTRODUCTION

*Helicobacter pylori (H. pylori) is* a well-defined, spiral-shaped, gram-negative bacterium which is the primary causative agent of several gastric pathologies; ranging from mild gastritis to gastric malignancies (Crowe, 2019). Infection with *H. pylori* causes chronic inflammation of the gastric mucosa and significantly increases the risk of developing duodenal and gastric ulcers and gastric cancer. *H. pylori* is responsible for 90-100% of duodenal ulcers and 70-90% of gastric ulcers cases (Karim et al., 2013) and infects approximately 50% of the world’s population (Rahman et al., 2013). The prevalence of infection varies widely depending on geographical area, age, race, ethnicity, and socioeconomic status. Rates appear to be higher in developing countries than in developed countries, with most infections occurring during childhood. The prevalence among middle-aged adults is over 80% in many developing countries, compared to 20 to 50 precents in industrialized countries (Suerbaum and Michetti, 2002). Infection persists throughout the life unless treated (Zali, 2011). Noteworthy to mention that healthy hygiene practices have been found to be associated with a decreased rate of *H. pylori* infections (Brown, 2000).

Similar to other developing countries, the rate of *H. pylori* infection is notably high in Bangladesh. In 1997, Ahmad et al. reported that the prevalence of *H. pylori* infection in Bangladesh was 92% in their serology-based study (Ahmad et al., 1997). Mahalanabis et al., in a study based on the 13C-urea breath test, also reported that the prevalence of *H. pylori* was 84% in 6-9 years old (Mahalanabis et al., 1996). The overall *H. pylori* prevalence in other Asian countries, including India (79% by ELISA) and Pakistan (84% by PCR), was also reported high (Ahmad et al., 1997; Graham et al., 1991). In Europe (40%) and the United States (40%), a significantly lower prevalence rate of *H. pylori* was observed (Andersen et al., 1996; Everhart et al., 2000). High infection rates in developing countries may be the consequence of poor socioeconomic conditions and unhygienic lifestyles (Rahman et al., 2013).

The association between *H. pylori* and gastroduodenal diseases necessitates the proper and timely diagnosis of infection in dyspeptic patients. Two broad methods are used to detect *H. pylori* in routine clinical diagnosis: invasive and non-invasive methods (Sabbagh et al., 2019). The methods that require endoscopy for examining the gastric mucosa are called invasive (direct) methods. Several tests can be performed on the gastric mucosa obtained by endoscopy: Rapid urease test (RUT), histology, microscopic examination of stained sμears, culture, and polymerase chain reaction (Sabbagh et al., 2019). Non-invasive methods, which do not necessitate an endoscopic examination, are - serology, urea breath test (UBT), and *H. pylori* stool antigen test (HpSAg) (Braden, 2012). Each of the aforementioned assays χoμes with underlying advantages and disadvantages and often, owing to undermined sensitivity and specificity, none of these assays can be considered as a gold standard.

Serology is widely used for screening patients for *H. pylori* infection. It is quick, relatively inexpensive, and has good sensitivity. However, Anti - *H. pylori* IgG can usually be detected by 3-4 weeks after infection, which is a drawback since this mode of testing is ineffective in detecting the presence of *H. pylori* during an earlier stage of pathogenesis (Moayyedi et al., 2003). An Anti - *H. pylori* IgG biochemical assay returns positive test results of the antibody secretion even after the bacterium has been eradicated. Hence, serology test results can be misleading and serve as false positive test results which could be invalid (Miftahussurur and Yamaoka, 2016). The urea breath test provides a reliable non-invasive method for the detection of *H. pylori* infection. It has high accuracy and reproducibility with sensitivity and specificity of 90-96% and 88-98%, respectively (Howden and Hunt, 1998). Nevertheless, it is relatively expensive and requires μass spectrometric analysis which may not be available in limited resource centers (Ansari and Yamaoka, 2018).

*H. pylori* also appear in stool as it is a gastrointestinal pathogen. Stool tests have the advantage of being non-invasive, and the specimen is easily obtainable. The monoclonal antibody-based *H. pylori* stool antigen (HpSAg) assay has been clinically useful with sensitivity and specificity of more than 90%. The specimen is easily obtainable and can be used as a routine diagnostic tool for *H. pylori* infection because it seems to overcome the conventional invasive techniques’ limitations (Ahmad et al., 1997; Khatoon et al., 2016). Detection of *H. pylori* antigen in fecal samples may be useful for non-invasive diagnosis of infection in both children and adults, selecting cases requiring endoscopic examination, and in epidemiological studies (Gulcan et al., 2005).

This present study was designed to analyze the diagnostic value of a rapid immunochromatography-based stool antigen test compared to the invasive tests – rapid urease test and histopathological analysis of gastric mucosal biopsy to diagnose *H. pylori* infection in adult patients with peptic ulcer disease before treatment. For a developing country like Bangladesh, the accessibility and reliability of rapid immunochromatography-based stool antigen tests can be useful in quicker detection of *H. pylori* infection, obtaining valid results during and post infection, in overcoming the economic and technical barriers of other biochemical assays and facilitate in proper disease management.

## 2. MATERIALS AND METHODS

### 2.1 Sample collection

A cross-sectional descriptive study was conducted in the Department of Microbiology, Department of Pathology and Department of Gastroenterology, Chattogram, Medical College and Hospital (CMCH), Chattogram over a period from July 2017 to June 2018 for a total of 140 patients with suspected peptic ulcer disease and 20 asymptomatic individuals of either sex from 18 to 70 years of age. To study a representative sample of the healthy population with active infection, stool samples from 20 healthy individuals were included in this group for HpSAg test. All of them were free from upper GI symptoms and did not undergo an endoscopic examination. The specimens were collected from the Department of Gastroenterology of Chattogram Medical College Hospital (CMCH). A structured questionnaire was developed to collect data from the patient. All patients were informed about the objective of the study. Every ethical issue was discussed with the patients regarding the study and informed written consent was obtained subsequently.

### 2.2 Inclusion and Exclusion criteria

Patients with symptoms related to the upper GI tract and symptoms of PUD (burning epigastric pain exacerbated by fasting and improved with meals, anorexia, nausea, vomiting, bloating, belching, etc.), diagnosed clinically, advised by physicians for endoscopic examination, and subsequently diagnosed with gastritis and peptic ulcer by endoscopic finding were included in this study.

Patients with other comorbid conditions who were not fit for endoscopy were not included in this study. Patients with a history of antimicrobial drugs, proton pump inhibitors, H2 receptor blockers, and bismuth preparation in the preceding two weeks or who received *H. pylori* eradication therapy in the previous six months were excluded. Moreover, patients with steroid therapy and other immunosuppressive drugs, patients with complicated peptic ulcers including active bleeding, perforation, and pyloric stenosis, and patients with gastric carcinoma were also excluded from the study.

Culture is usually considered the gold standard for the diagnosis of any microorganism. However, in the current study, culture was not carried out because of some drawbacks, including that the microorganism is slow-growing, fastidious. The ability to isolate the organism from infected subjects varies widely between laboratories and makes it the most technically demanding *H. pylori* diagnostic test (Waidyarthne et al., 2012). Therefore, to determine the HpSAg, stool samples were collected from 20 healthy individuals with no history of dyspepsia, peptic ulcer disease, and anti-H. *pylori* therapy in the previous six months. These individuals were considered as disease negative or “Healthy Controls”.

### 2.3 Collection and experiments of specimens

#### 2.3.1 Gastric biopsy

Selected patients underwent upper GI endoscopy in the Department of Gastroenterology, CMCH by expert endoscopists for possible detection of peptic ulcer disease. A total of 140 patients who fulfilled the inclusion criteria attending the gastroenterology outpatient department, were initially enrolled for upper GI endoscopy. Patients found to have ulcers or erosions anywhere in the stomach or duodenums up to the second part of endoscopy were selected to take a biopsy. A total of 75 patients with endoscopic findings of peptic ulcer disease were finally included in the study.

Ulcer was diagnosed at endoscopy in the stomach or duodenum when there was a mucosal break of diameter 5 mm or larger, covered with fibrin. Mucosal breaks smaller than 5mm were considered as erosions (Alam et al., 2014). Presence of lesions in the gastroduodenal mucosa was noted. Biopsy specimens were sampled from both gastric antrum and corpus. Three biopsy tissues were obtained from the gastric antrum and one from the upper corpus of the stomach of each patient. Two biopsy specimens of the antrum and one of the corpus will be fixed in 10% buffered formalin for histopathology. The other biopsy specimen from the antrum was incubated in Rapid urease test (RUT) kit and the results were recorded in the data sheet.

#### 2.3.2 Laboratory procedures

RUT was performed instantly on one antral biopsy specimen using a χoμμerχially available kit Pronto Dry. Results were available within a few minutes to a few hours and always within 24 hours (Alam et al., 2014). Histopathological examination was performed from both antrum and gastric corpus biopsy samples. Biopsy specimens for histopathology were processed according to the standard procedure described before. This procedure was done in the Department of Pathology, CMC, with the samples supplied. In microscopic examination, the organism can be readily recognized by its characteristic curved or ‘S’ - shaped morphology and its location on the epithelial cell surface, within the gastric pits or in the mucus gel overlying the cell surface.

#### 2.3.3 Immunochromatographic test (ICT) of stool for *H. pylori* antigen

Patients were asked to collect a specimen from their first stool sample after endoscopy. HpSAg test was done in the 75 patients and 20 healthy controls. The H. pylori antigen rapid test cassette (faeces), is a commercially available, rapid chromatographic immunoassay for the qualitative detection of H. pylori antigen in human faeces. Detail procedure has been described in previous sections.

### 2.4 Data analysis

All relevant medical history, physical examination records, clinical findings, and laboratory records of every subject were systematically added in a predesigned datasheet for subsequent analysis. The data were checked and edited after collection. Continuous variables were reported as the mean ± SD, and categorical variables were reported as percentages. Two tests for categorical data compared baseline characteristics. Statistical significance was defined as p <0.05, and the confidence interval was set at the 95% level. SPSS (Statistical Package for Social Science) version 20 software was used for the analyses.

## 3. RESULTS

### 3.1 Clinical characteristics of the patients

Table 1 represents the age distribution of the patients with dyspepsia based on the endoscopic diagnosis. Among the 140 patients, 77 (55.0%) were in the 21 to 40 years age group, 48 (34.3%) belonged to the 41 to 60 years age group, followed by 11 (7.8%) in the age group above 60 years and 4 (2.86%) patients were in the age group below 20 years. Endoscopic diagnosis revealed that most of the patients (55.0%) with dyspepsia were in the age group of 21 to 40 years, followed by 48 (34.3%) in the age group 41 to 60 below 20 years were less. The mean age of the patients was 39.4±11.4. Of the study participants, 76 (54.3%) were male, and 64 (45.7%) were female. The male: female ratio was found to be1.06: 1. No association was found between the endoscopic findings and the gender of the patients

**Table 1:** Association of *H. pylori* infection prevalence with age and gender.

| Characteristics | N=140 (%) | Ulcer & erosion; n=75 (53.6%) | No ulcer/erosion; n=65 (46.4%) | Chi-square | P-value |
| --- | --- | --- | --- | --- | --- |
| Age (years); Mean age= 39.4±11.4 (SD) |  |  |  |  |  |
| <20 | 4 (2.9%) | 2 (50.0%) | 2 (50.0%) | 15.4 | 0.0015 |
| 21-40 | 77 (55.0%) | 52 (67.5%) | 25 (32.5%) |  |  |
| 41-60 | 48 (34.3%) | 19 (39.6%) | 29 (60.4%) |  |  |
| >60 | 11 (7.8%) | 2 (18.2%) | 9 (81.8%) |  |  |
| Gender; male: female ratio of 1.06:1 |  |  |  |  |  |
| Male | 76 (54.3%) | 43 (56.6%) | 33 (43.4%) | 0.6 | 0.4368 |
| Female | 64 (45.7%) | 32 (50.0%) | 32 (50.0%) |  |  |

SD, Standard deviation

Among the different categories of peptic ulcer, antral erosion (25.7%) and prepyloric erosion (15.0%) were more prevalent (Figure 1A) (Supplementary figure 1). The proportion of antral erosion was 57.4% in patients who were below 40 years. This particular type of ulcer decreased to 23.4% in patients over 40 years. Patients over 40 years were mostly suffering from Prepyloric erosion (42.9%) (Figure 1B). The non-ulcer patients’ percentage of reflux oesophagitis increased by about 10% in patients over 40 years compared with patients below 40 years (Figure 1C).

**Figure 1:**
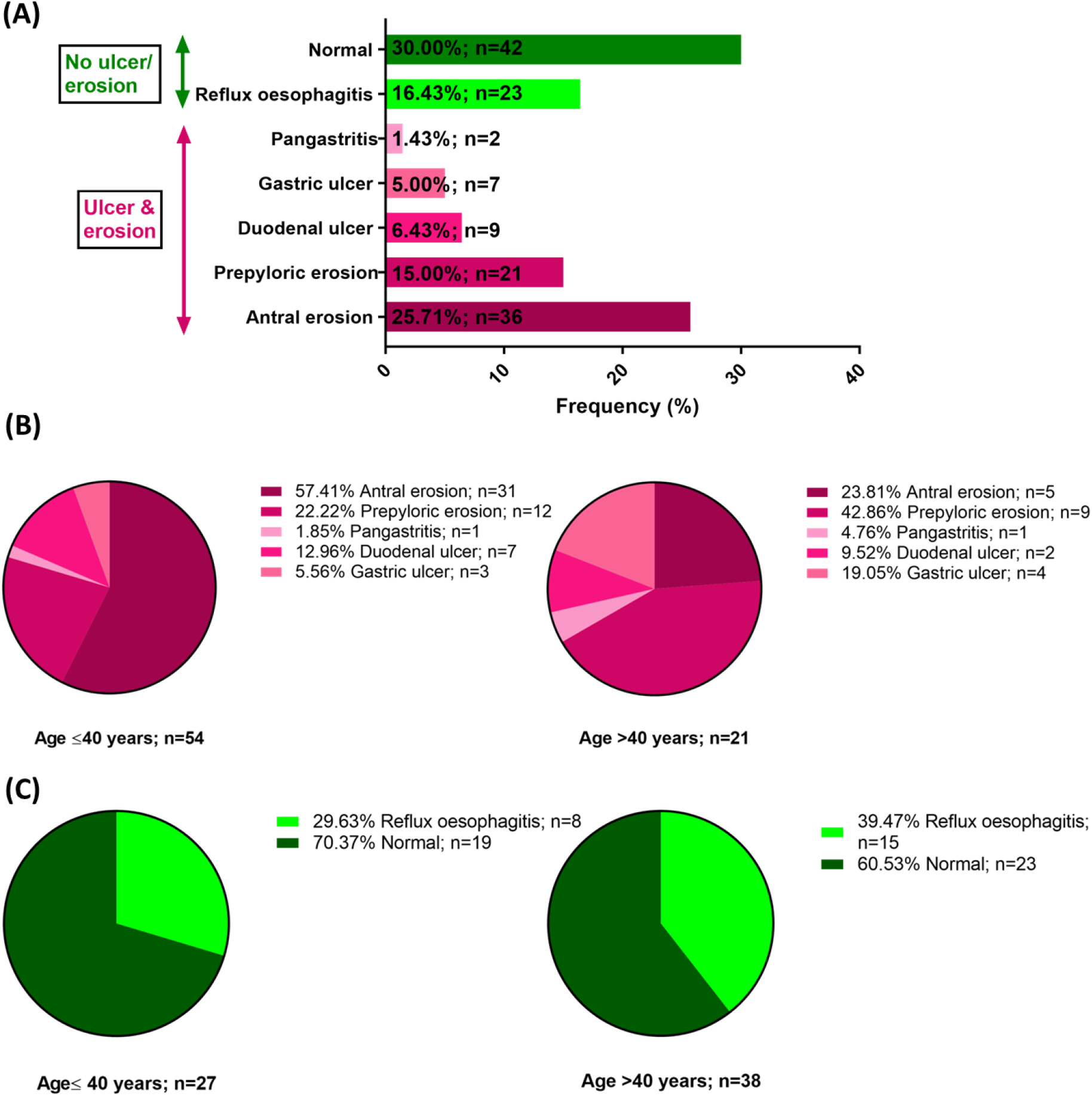
Identification and prevalence of peptic ulcer in patients with dyspepsia based on endoscopic diagnosis. (A) Prevalence of different types peptic ulcers in the studied patients; (B) Frequency of different type peptic ulcers in patients below and over 40 years; (C) Frequency of normal and reflux oesophagitis cases in non-peptic ulcer patients below and over 40 years.

### 3.2 Different *H. pylori* detection approaches to define *H. pylori* positive patients

Comparative results of all four techniques unitized in this study to detect *H. pylori* have been illustrated in Figure 2. Histopathological grading of chronic gastritis identified the presence of *H. pylori* in a total of 75 patients. However, microscopic grading of curved bacilli of *H. pylori* was identified in 70 patients. Among these 70 patients, 67 were *H. pylori positive* in the rapid urease test (RUT), and 65 were *H. pylori positive* in HpSAg test.

**Figure 2:**
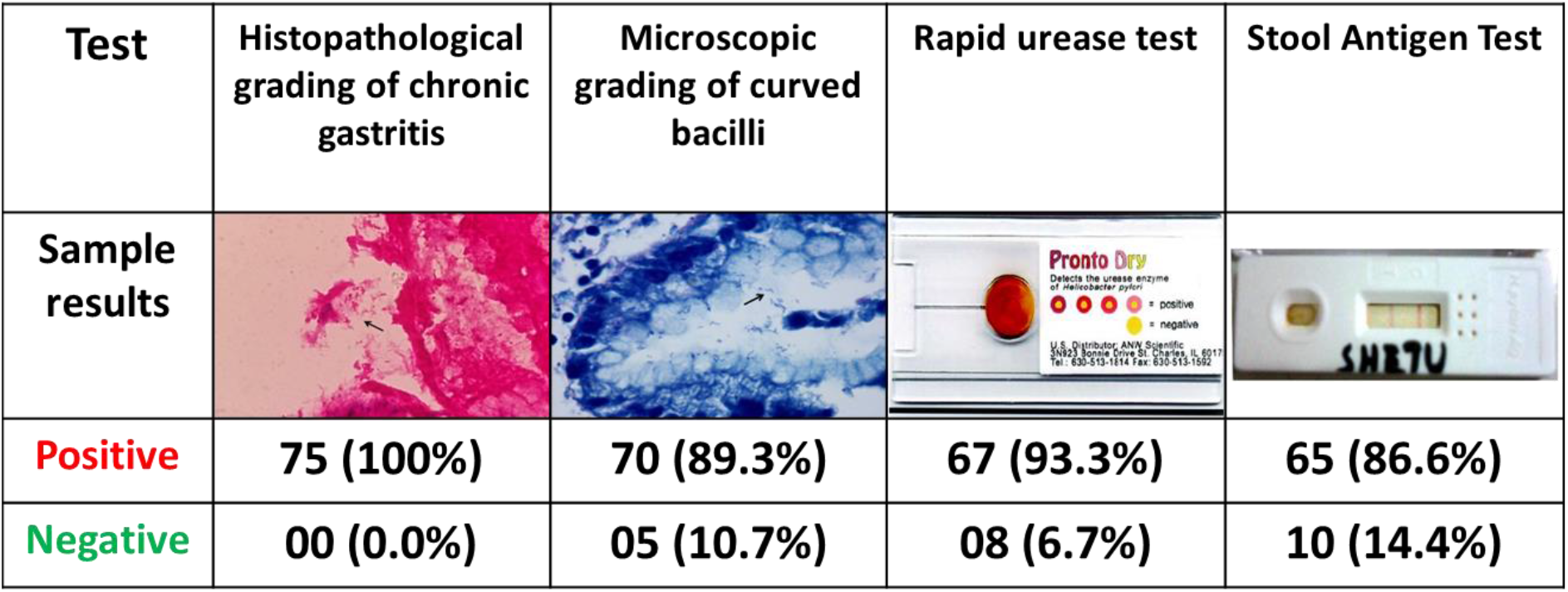
All four procedures were used to identify *H. pylori* in the studied subjects.

Figure 3A shows the histopathologiχal grading of chronic gastritis. Out of 75 patients, 59 (78.0%) had mild inflammation, 14 (18.7%) had moderate inflammation, and 2 (2.7%) had chronic active gastritis. Figure 3B shows the baχterial density and severity of gastritis among the studied subjects. The highest percentage of patients was in grade 2 (44.0%), and the next highest was in the grade 1 group (36.0%). The rest of the patients who were found to have bacilli were in group 3 (13.3%). Bacilli were found in a total of 70 (93.3%) cases. Figure 3C and Table 2 show the rapid urease test (RUT) results in endoscopically proved ulcer and erosion cases. Among 75 patients, 67 (89.3%) were RUT positive, and 8 (10.7%) were RUT negative.

**Table 2:** Demonstration of curved bacilli on microscopic examination & its relation to Histopathology and Rapid urease test [n=75].

|  | No. Of cases | Microscopic grading of curved bacilli |  |  | No Bacilli seen |
| --- | --- | --- | --- | --- | --- |
|  |  | Grade 1 | Grade 2 | Grade 3 | Grade 0 |
| Histopathological grading of chronic gastritis |  |  |  |  |  |
| Mild | 59 | 25 | 23 | 6 | 5 |
| Moderate | 14 | 2 | 9 | 3 | 0 |
| Chronic active | 2 | 0 | 1 | 1 | 0 |
| Total | 75 | 27 | 33 | 10 | 5 |
| Rapid urease test (RUT) |  |  |  |  |  |
| Positive | 67 | 24 | 33 | 10 | 0 |
| Negative | 8 | 3 | 0 | 0 | 5 |
| Total | 75 | 27 | 33 | 10 | 5 |

RUT, Rapid urease test

**Figure 3:**
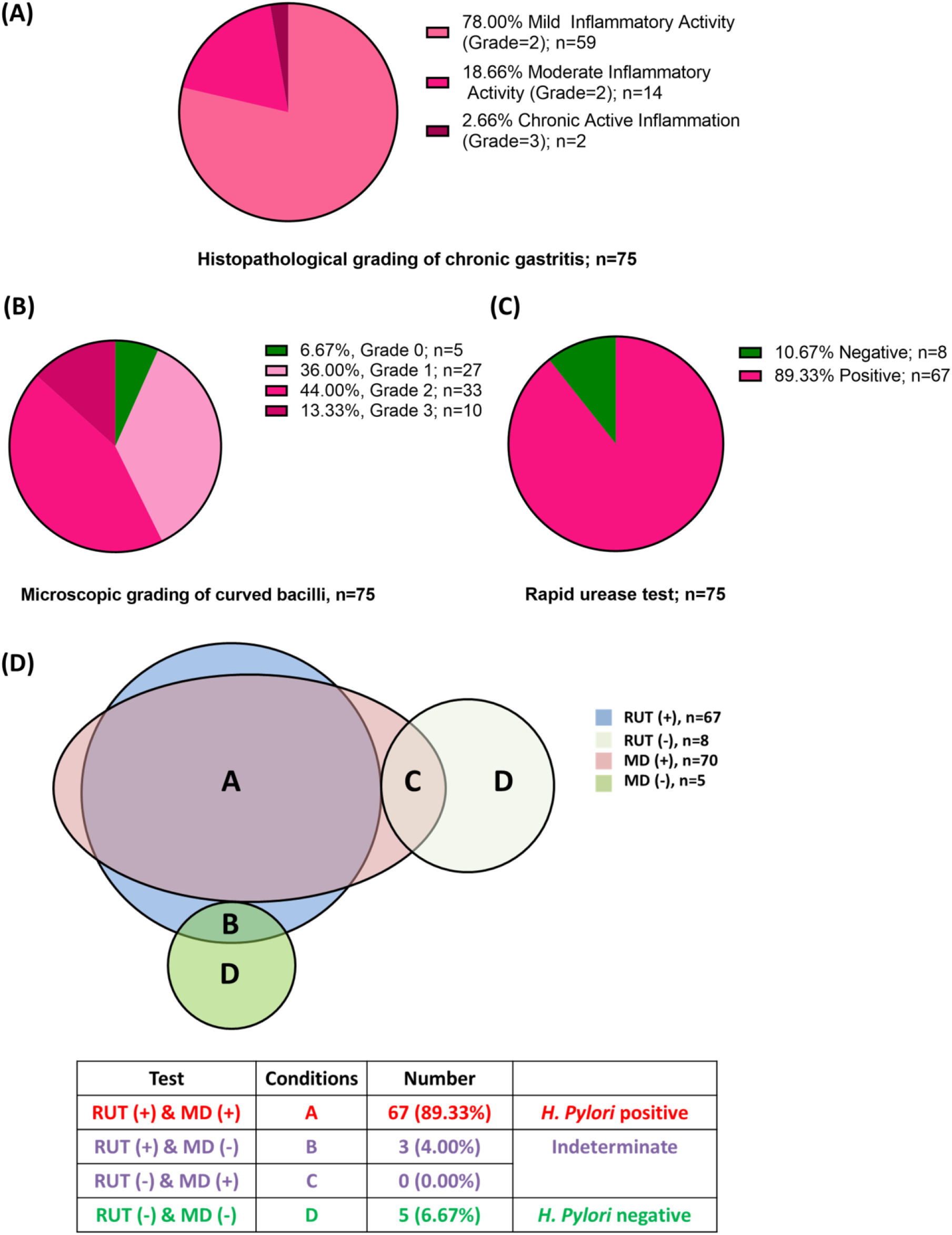
Different approaches to identify *H. pylori* status in patients. (A) Frequency of histopathological grading of chronic gastritis; (B) Microscopic detection (MD) of curved bacilli in endoscopically proved ulcer and erosion cases; (C) Rapid urease test (RUT) result to identify *H. pylori* positive patients in endoscopically proved ulcer and erosion cases; (D) Comparative result of microscopic detection (MD) and rapid urease test (RUT) to define *H. pylori* positive patients. Either RUT or microscopy positive but not both positive were defined as indeterminate states. Three cases were microscopy positive but RUT negative.

Table 2 shows the relationship between bacterial density and severity of gastritis. Out of the 59 cases with mild inflammation, the bacterial density was as follows: 25 showed grade – 1 bacterial load, 23 had grade – 2, 6 patients showed grade – 3 load, but no bacilli were seen in 5 cases. Among the 14 moderate inflammation cases, 2 had grade – 1, 9 had grade – 2, and 3 had grade – 3 bacterial load. Among the two cases of chronic active inflammation, 1 showed grade – 2 and 1 showed grade – 3 bacterial density. Interestingly, among the eight RUT negative patients, three had grade 1 microscopic curved bacilli (Table 2).

Figure 3D shows the combined result of RUT and microscopic grading detection (MD) of curved bacilli, the *H. pylori* status of the patient. According to the definition of the gold standard, both invasive test positive cases were considered *H. pylori-positive* states, and both invasive test negative cases were considered *H. pylori-negative* states. Any one of the test’s positive cases was considered as indeterminate. Out of 75 cases, 67 were *H. pylori-positive*, 5 cases were *H. pylori-negative*, and 3 cases were indeterminate. Either RUT or MD positive, but not both positive, was defined as an indeterminate state. All three indeterminate cases were MD positive but RUT negative.

### 3.3. Sensitivity of *H. pylori* stool antigen test (HpSAg)

Table 3 represents the stool antigen detection results among *H. pylori-positive* and negative cases. Out of 67 *H. pylori-positive* cases, 64 (95.52%) were positive in HpSAg test. The remaining 3 (4.48%) cases showed false-negative results. The HpSAg test was negative in all 5 (100%) of *H. pylori-negative* status patients. No false-positive results were obtained from these patients. Among the three indeterminate cases, 1 (33.33%) was positive for HpSAg test, and 2 (66.67%) were negative. A strong association was found between *H. pylori* status and positivity or negativity of HpSAg test. *H. pylori-positive* status patients showed more positive HpSAg test results than the H. pylori-negative and indeterminate status group.

**Table 3:**
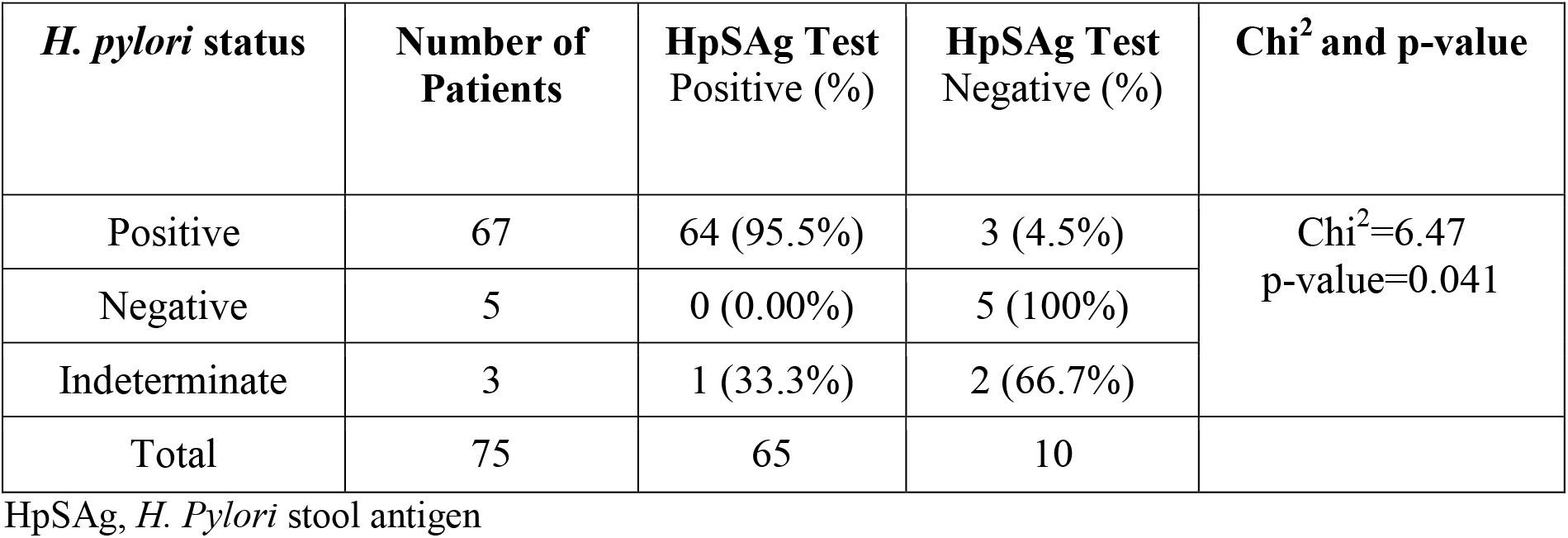
HpSAg detection results among the *H. pylori* positive & negative cases [n=75].

| *H. pylori* status | Number of Patients | HpSAg Test Positive (%) | HpSAg Test Negative (%) | Chi<sup>2</sup> and p-value |
| --- | --- | --- | --- | --- |
| Positive | 67 | 64 (95.5%) | 3 (4.5%) | Chi<sup>2</sup>=6.47<br>p-value=0.041 |
| Negative | 5 | 0 (0.00%) | 5 (100%) |  |
| Indeterminate | 3 | 1 (33.3%) | 2 (66.7%) |  |
| Total | 75 | 65 | 10 |  |

HpSAg, *H. Pylori* stool antigen

Performance of HpSAg test is presented in Table 4. To determine the specificity of the HpSAg test, stool samples were collected from 20 healthy individuals with no history of dyspepsia, peptic ulcer disease and anti *H. pylori* therapy in the previous 6 months. Among the 20 healthy controls, 16 (80%) were negative and 4 (20%) cases were positive for HpSAg test. The study found that the immunochromatographic test for HpSAg detection is highly sensitive and specific; sensitivity 95%, specificity 80%, Positive predictive value (PPV) 94%, Negative predictive value (NPV) 84% and diagnostic accuracy 91%.

**Table 4:** Evaluation of HpSAg detection results in *H. pylori* positive cases & χontrols [n=67+20].

| HpSAg Test | Case | Control | Total |  |
| --- | --- | --- | --- | --- |
| Positive | 64 | 4 | 68 | **Sensitivity of stool ICT : 95%**<br>**Specificity of stool ICT : 80%**<br>**PPV of stool ICT : 94%**<br>**NPV of stool ICT : 84%**<br>**Diagnostic Accuracy : 91%** |
| Negative | 3 | 16 | 19 |  |
| Total | 67 | 20 | 87 |  |

HpSAg, *H. Pylori* stool antigen; ICT, Immunochromatographic Test; NPV, Negative predictive value; PPV, Positive predictive value;

## 4. DISCUSSION

In this study, out of 140 patients with upper GIT symptoms, the highest incidence (55%) was observed in the 21-40 years age group, followed by 41-60 years (34.28%) age group (Table 1). The patients’ age ranged from 18 to 70 years, with a mean age of 39.38 years. A similar result was reported by Islam et al. (2010) in Dhaka, Bangladesh, where 60% of cases were within the 21-40 years age group, followed by 27% cases in the 41-60 years age group (Islam, 2010). The mean age of the patients was 37.98 years. Moreover, the proportion of male and female patients was 51.43% and 48.57%, respectively, with only a slight predominance of male patients showing a male: female ratio of 1.06:1 (Table 1). This finding is in harmony with the study done in Turkey by (Korkmaz et al., 2015), where 49.4% and 50.6% of the study subjects were male and female, respectively.

Overall findings of endoscopy showed 30% normal, 16.43% reflux oesophagitis, 25.71% antral erosion, 15% erosion in body and fundus, 5% gastric ulcer, and 6.43% duodenal ulcer (Figure 1A). These findings are similar to those of Ghosh et al. (2013), in Dhaka, Bangladesh, where among 72 dyspeptic patients, gastro-duodenal mucosa was normal in 55.6%, reflux oesophagitis in 8.3%, gastric erosion in 22.2%, gastric ulcer in 2.8% and duodenal ulcer in 5.7% of patients (Ghosh et al., 2017). However, our finding differs from the study of Ayana et al. (2014), done in Tanzania, where 94.2% of the subjects had abnormal findings and only 5.8% were found to be expected in endoscopy (Ayana et al., 2014). There was gastritis in 61.1% of patients, gastroesophageal reflux disease (GERD) in 58.7% of patients, and peptic ulcer disease (duodenal and gastric ulcers) in 24.1% of patients. The differences may be attributed to the exclusion and inclusion criteria used in different studies, socioeconomic status differences, and attitude towards seeking healthcare.

As for histopathology, chronic inflammatory infiltrates were present in almost all cases, with the majority (78%) having only mild inflammation (Figure 3A). The density of *H. pylori* colonization increased with the severity of chronic inflammation in our study (Table-2). These findings were compatible with the study performed by Garg et al. (2012) in India, who reported that 70% of cases had mild inflammation (Garg et al., 2012). A greater density of organism was noted in higher grades of inflammation. However, Priyadarshini et al. (2018) found moderate inflammation in most cases (58.2%) (Priyadarshini M. M., 2018). In contrast to the present study, Park et al. (1995), in Seoul, Korea, found that there was marked variation between the intensity of inflammation and the number of organisms in a histologic section (Park et al., 1995). *H. pylori* concentration did not affect the activity and overall histological grading of chronic gastritis in their study. This dissimilarity may be due to genetic differences, nutritional habits, and environmental factors between the two study populations (Ghasemi et al., 2017). Among the 75 patients, 67 (89.33%) were positive, and 8 (10.67%) were found to be negative for the rapid urease test (RUT) in our study (Figure 3C). Alam et al. (2014) reported 88.76% RUT positivity among PUD patients in Dhaka. These findings are consistent with that of the present study (Alam et al., 2014). In contrast to the present study, Islam et al. (2013), reported lower RUT positivity (61.7% of patients were RUT positive and 38.3% were RUT negative) . This dissimilarity may be because sensitivity can vary with the site chosen for biopsy due to the bacteria’s patchy distribution; hence, a false negative test can occur due to sampling error (Waidyarthne et al., 2012).

In this study, 67 of the 75 cases were rapid urease tests and histopathologically positive for *H. pylori* (Figure 3C). As a result, 67 (89.33%) patients were considered infected with *H. pylori*. This is in accordance with the study performed by Islam et al. (2010) and Alam et al. (2014) in Bangladesh, who reported 79.02% and 87.64% cases were infected by *H. pylori*, using the same definition (Alam et al., 2014; Islam, 2010). This finding is in contrast with the lower detection rate of (50.6%) *H. pylori-positive* cases by Korkmaz et al. (2015) in Turkey (Korkmaz et al., 2015). The higher detection rate is not abnormal as the study was conducted in a developing country where 80% - 90% of the population are estimated to be carriers of this pathogen. The lower detection rate in the above study was probably due to the study being done in a developed country.

To determine the specificity of the HpSAg test, stool samples were collected from 20 healthy individuals with no history of dyspepsia, peptic ulcer disease, and anti-H. *pylori* therapy in the previous six months. These individuals were considered as disease negative or “Healthy Controls”. Of the 20 control stool samples, 4 (20%) cases were positive for the HpSAg test (Table 4). A strong association was found between the presence of *H. pylori* and positivity or negativity of stool antigen test. *H. pylori-positive* status patients were given more positive HpSAg tests. On the other hand, disease-negative healthy control subjects were found to have more negative HpSAg tests than positive ones. Our study’s most important finding was that the new HpSAg test showed excellent sensitivity and specificity. The sensitivity, specificity, PPV, NPV, and diagnostic accuracy of the HpSAg test were 95%, 80%, 94%, 84%, and 91%, respectively (Table 4).

Several studies on monoclonal stool antigen tests based on immunochromatography in various regions of the world have shown comparable results. A similar study done in Malaysia showed sensitivity, specificity, PPV, NPV, and accuracy as 91.7%, 100%, 100%, 94.6%, and 96.6%, respectively (Osman et al., 2014). In addition to this, a study done in Korea found the sensitivity, specificity, PPV, NPV, and accuracy of *H. pylori* stool immunochromatographic antigen assay (S-ICT test) as 84.5%, 96.2%, 95.6%, 86.4%, and 90.4%, respectively (Jekarl et al., 2013). However, these results differ from the findings of a study performed in Turkey by Korkmaz et al. (2015) (Korkmaz et al., 2015). They reported the sensitivity, specificity, PPV, NPV, and accuracy of an immunochromatographic HpSAg test as 51.6%, 96%, 88.8%, 76.1% and 79%, respectively. Lower sensitivities for HpSAg tests have occurred in special circumstances, such as those undergoing PPI or bismuth therapy, patients with liver cirrhosis, or hidden gastrointestinal bleeding (Jekarl et al., 2013), which may have resulted in false-negative HpSAg test. Nonetheless, the low colonization of bacteria in the stomach and the consequent low concentrations of *H. pylori* antigen in the feces could be sufficient to cause false-negative results (Korkmaz et al., 2015).

The present study’s data may not be a precise estimate of the prevalence of *H. pylori* infection in Chittagong because the sample we studied was not adequately large. Hence, further large-scale studies are required to for a more transparent understanding of the epidemiology of *H. pylori* infection and its association with disease outcomes in this region and other regions of Bangladesh. To the best of our knowledge, this is the first prospective study to determine the efficacy of a HpSAg test for the diagnosis of infection in Chittagong Medical College, the biggest government hospital in Bangladesh’s southern region. Therefore, this study will play a significant role in gaining acceptance of the HpSAg test among doctors and patients in Bangladesh’s greater Chattogram region as a simple and inexpensive process.

## 5. CONCLUSION

*H. pylori* stool ICT is a rapid, easy, non-invasive, and inexpensive method for detecting infection. This test showed high sensitivity and specificity. No significant differences were found in sensitivity or specificity between the gold standard (invasive) tests (RUT and microscopic examination of gastric tissue) and the HpSAg test. Therefore HpSAg test can be a reliable alternative to other techniques to diagnose active *H. pylori* infection. This test can also be used in some special conditions, for example, if endoscopy is not available. HpSAg test is a sustainable alternative to invasive tests for the detection of *H. pylori* infection in children, especially in developing countries like Bangladesh where the prevalence of infection is very high. The test may further be used in monitoring the therapeutic response in *H. pylori* infection.

## Data Availability

All data produced in the present study are available upon reasonable request to the authors

---

**Supplementary figure 1:**
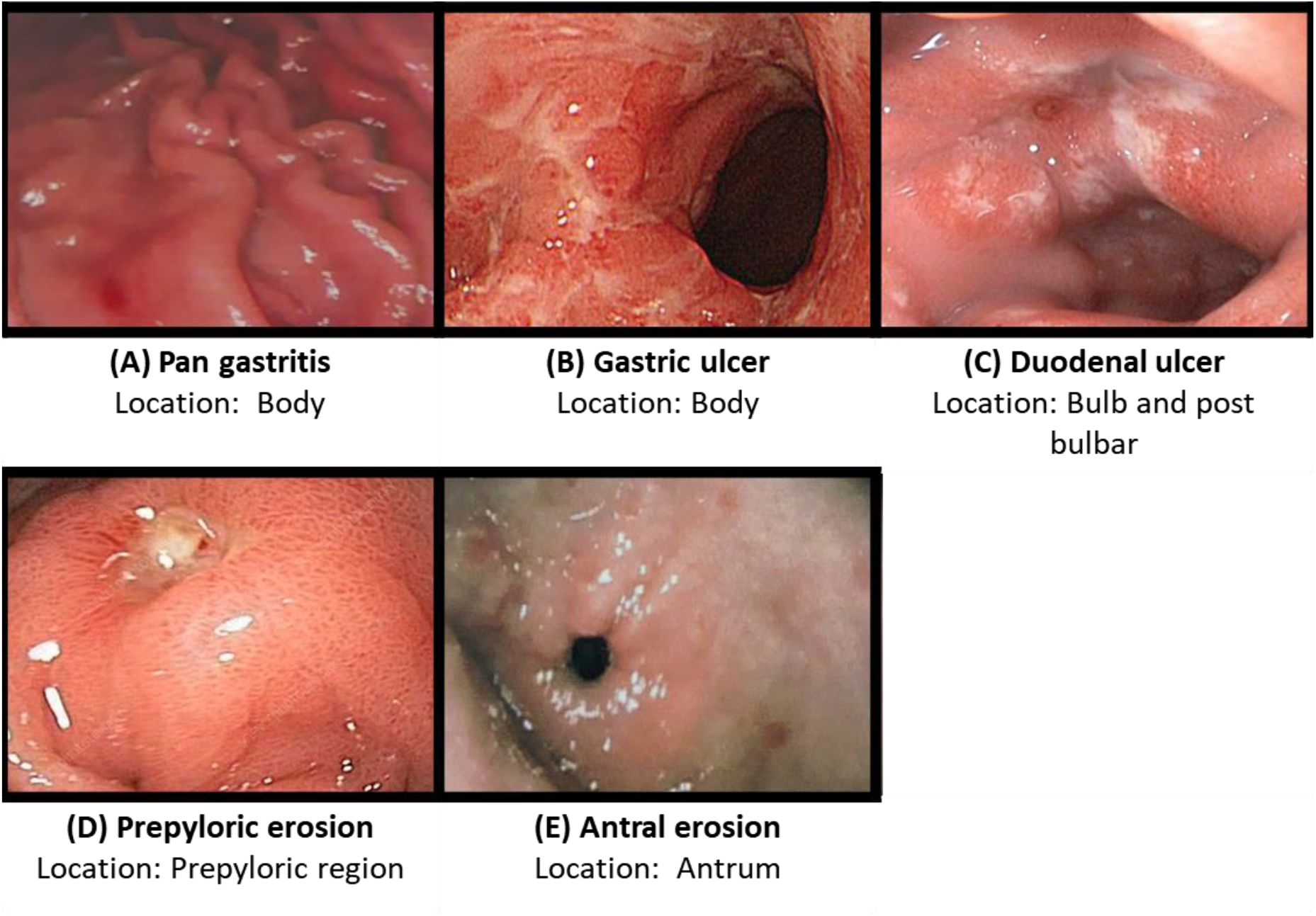
Types of ulcer and erosion identified on endoscopy examination. (A) Pan gastritis iμages taken from the body area of the stomach; (B) Gastric ulcer image taken from the body area of the stomach; (C) Duodenal ulcer image taken from Bulb and post-bulbar areas; (D) Prepyloric erosion image taken from prepyloric region; (E) Antral erosion image taken from antrum area.

## REFERENCES

Ahmad, M.M., Rahman, M., Rumi, A.K., Islam, S., Huq, F., Chowdhury, M.F., Jinnah, F., Morshed, M.G., Hassan, M.S., Khan, A.K., et al. (1997). Prevalence of Helicobacter pylori in asymptomatic population--a pilot serological study in Bangladesh. Journal of epidemiology 7, 251–254.

Alam, M.R., Ahmed, E., Rahman, M.Z., Islam, A., Khan, M.M., Ahmed, D., Masud, H., and Raihan, A. (2014). A study on healing of peptic ulcer disease after eradication of Helicobacter pylori infection. Bangladesh Medical Journal 43, 84–89.

Andersen, L.P., Rosenstock, S.J., Bonnevie, O., and Jørgensen, T. (1996). Seroprevalence of immunoglobulin G, M, and A antibodies to Helicobacter pylori in an unselected Danish population. American journal of epidemiology 143, 1157–1164.

Ansari, S., and Yamaoka, Y. (2018). Current understanding and management of Helicobacter pylori infection: an updated appraisal. F1000Research 7.

Ayana, S.M., Swai, B., Maro, V.P., and Kibiki, G.S. (2014). Upper gastrointestinal endoscopic findings and prevalence of Helicobacter pylori infection among adult patients with dyspepsia in northern Tanzania. Tanzania journal of health research 16, 16–22.

Braden, B. (2012). Diagnosis of Helicobacter pylori infection. BMJ (Clinical research ed) 344, e828.

Brown, L.M. (2000). Helicobacter pylori: epidemiology and routes of transmission. Epidemiologic reviews 22, 283–297.

Crowe, S.E. (2019). Helicobacter pylori Infection. The New England journal of medicine 380, 1158–1165.

Everhart, J.E., Kruszon-Moran, D., Perez-Perez, G.I., Tralka, T.S., and McQuillan, G. (2000). Seroprevalence and ethnic differences in Helicobacter pylori infection among adults in the United States. The Journal of infectious diseases 181, 1359–1363.

Garg, B., Sandhu, V., Sood, N., Sood, A., and Malhotra, V. (2012). Histopathological analysis of chronic gastritis and correlation of pathological features with each other and with endoscopic findings. Polish journal of pathology : official journal of the Polish Society of Pathologists 63, 172–178.

Ghasemi, B., Hamid, R., Ghobakhlou, M., Akbari, P., Dehghan Arash, Seif Rabiei, and Mohamad Ali (2017). Correlation between the intensity of Helicobacter pylori colonization and severity of gastritis. Gastroenterology Research and Practice 2017, 8320496.

Ghosh, C.K., Khan, M.R., Alam, F., Shil, B.C., Kabir, M.S., Mahmuduzzaman, M., Das, S.C., Masud, H., and Roy, P.K. (2017). Peptic ulcer disease in Bangladesh: A multi-centre study. Mymensingh medical journal : MMJ 26, 141–144.

Graham, D.Y., Adam, E., Reddy, G.T., Agarwal, J.P., Agarwal, R., Evans, D.J., Jr., Malaty, H.M., and Evans, D.G. (1991). Seroepidemiology of Helicobacter pylori infection in India. Comparison of developing and developed countries. Digestive diseases and sciences 36, 1084–1088.

Gulcan, E.M., Varol, A., Kutlu, T., Cullu, F., Erkan, T., Adal, E., Ulucakli, O., and Erdamar, S. (2005). Helicobacter pylori stool antigen test. The Indian Journal of Pediatrics 72, 675–678.

Howden, C.W., and Hunt, R.H. (1998). Guidelines for the management of Helicobacter pylori infection. Ad hoc committee on practice parameters of the american college of gastroenterology. The American journal of gastroenterology 93, 2330–2338.

Islam, M., Rahman, S., Shamsuzzaman, S., Muazzam, N., Kibria, S., Hossain, M., Ahmed, N., Sarkar, A., & Nahar, S. (2010). A comparative study among different invasive methods for the diagnosis of Helicobacter pylori. Faridpur Medical College Journal 5, 21–24.

Jekarl, D.W., An, Y.J., Lee, S., Lee, J., Kim, Y., Park, Y.J., Kim, T.J., Kim, J.I., and Park, S.H. (2013). Evaluation of a newly developed rapid stool antigen test using an immunochromatographic assay to detect Helicobacter pylori. Japanese journal of infectious diseases 66, 60–64.

Karim, R., Ahmed, S.M., and Begum, F. (2013). Non-invasive stool antigen test for screening of Helicobacter pylori infection and assessing efficacy of treatment in patients with peptic ulcer. South East Asia Journal of Public Health 2, 28–33.

Khatoon, J., Rai, R.P., and Prasad, K.N. (2016). Role of Helicobacter pylori in gastric cancer: Updates. World J Gastrointest Oncol 8, 147–158.

Korkmaz, H., Findik, D., Ugurluoglu, C., and Terzi, Y. (2015). Reliability of stool antigen tests: investigation of the diagnostic value of a new immunochromatographic Helicobacter pylori approach in dyspeptic patients. Asian Pacific journal of cancer prevention : APJCP 16, 657–660.

Mahalanabis, D., Rahman, M.M., Sarker, S.A., Bardhan, P.K., Hildebrand, P., Beglinger, C., and Gyr, K. (1996). Helicobacter pylori infection in the young in Bangladesh: prevalence, socioeconomic and nutritional aspects. International journal of epidemiology 25, 894–898.

Miftahussurur, M., and Yamaoka, Y. (2016). Diagnostic methods of Helicobacter pylori infection for epidemiological studies: Critical importance of indirect test validation. Biomed Res Int 2016, 4819423–4819423.

Moayyedi, P., Deeks, J., Talley, N.J., Delaney, B., and Forman, D. (2003). An update of the Cochrane systematic review of Helicobacter pylori eradication therapy in nonulcer dyspepsia: resolving the discrepancy between systematic reviews. The American journal of gastroenterology 98, 2621–2626.

Osman, H.A., Hasan, H., Suppian, R., Bahar, N., Hussin, N.S., Rahim, A.A., Hassan, S., Andee, D.Z., and Zilfalil, B.A. (2014). Evaluation of the Atlas Helicobacter pylori stool antigen test for diagnosis of infection in adult patients. Asian Pacific journal of cancer prevention : APJCP 15, 5245–5247.

Park, J., Kim, M.K., and Park, S.M. (1995). Influence of Helicobacter pylori colonization on histological grading of chronic gastritis in Korean patients with peptic ulcer. Korean J Intern Med 10, 125–129.

Priyadarshini M.M.G.M.M., Manjunatha Y. A (2018). Significance of histologic grading using visual analogue scale in chronic gastritis. Tropical Journal of Pathology & Microbiology 4, 82–87.

Rahman, M.M., Rowshon, A., and Rahim, S. (2013). Diagnosis of Helicobactor pylori in Bangladesh: Limited options and utility of serologic test. Journal of Shaheed Suhrawardy Medical College, 5, 1–2.

Sabbagh, P., Mohammadnia-Afrouzi, M., Javanian, M., Babazadeh, A., Koppolu, V., Vasigala, V.R., Nouri, H.R., and Ebrahimpour, S. (2019). Diagnostic methods for Helicobacter pylori infection: ideals, options, and limitations. European Journal of Clinical Microbiology & Infectious Diseases 38, 55–66.

Suerbaum, S., and Michetti, P. (2002). Helicobacter pylori infection. The New England journal of medicine 347, 1175–1186.

Waidyarthne, E., Mudduwa, L., Lekamwasam, J., and Lekamwasam, S. (2012). Helicobacter pylori detection techniques: comparison of sensitivity, specificity and cost. Galle Medical Journal 17, 1.

Zali, M.R. (2011). Facing resistance of H.pylori infection. Gastroenterol Hepatol Bed Bench 4, 3–11.

