## Supplementary figures and images for "Stool Antigen Test is Effective and Sensitive for Detecting *Helicobacter Pylori* Infection in Bangladeshi Peptic Ulcer Patients"

### Supplemental figure 1

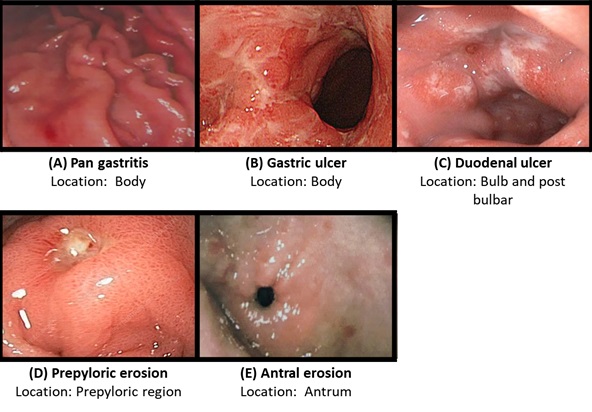
